# Age-structured dynamics and susceptibility in the face of infection and vaccination

**DOI:** 10.64898/2026.02.10.26345956

**Authors:** Ruiyun Li, Merawi Aragaw, Justin Maeda, C. Jessica E. Metcalf, Ottar N. Bjørnstad, Nils Chr. Stenseth

## Abstract

**Background:** Strikingly low allocation of SARS-CoV-2 vaccine to the African Continent limits its capacity to control transmission. Characterizing the trajectory of vaccination efforts and their impact on the expected burden of SARS-CoV-2 will help planning vaccine delivery strategies, and public health interventions more broadly. As the burden is strongly age-dependent, this requires an understanding of the age-structured dynamics of susceptible individuals, accounting for the combined effects of vaccination and infection induced immunity.

**Methods and Findings:** We illustrate with projections for diverse African LMIC demographics. To this end, we develop an age-structured mathematical model with vaccination to assess the likely time-horizon to reach target vaccine coverage of high-risk groups, and how susceptibility patterns across age will shift as a result of both infection, and the broadening of vaccination targets from a focus on risk groups to efforts to reach the general population. We base our assessment on the demography, contact patterns and public health capacity of 16 African countries with diverse age pyramids. We identify a considerable divergence in the projected horizon of expanded targeting from prioritized age groups to general vaccination, with longer time among those with higher mean age and lower vaccination capacity. We parameterize the model using realistic demographies and contact patterns to project the changing age profile of susceptibles. We demonstrate that contacts and vaccination jointly drive the early age profile; while immune duration contributes to the transition of age-susceptibility profile in the intermediate future.

**Conclusions:** Our model framework provides a flexible and critical preparedness-tools to inform decision making against future epidemic waves and beyond Covid-19.

## Introduction

The COVID-19 pandemic continues to ravage the global community with concurrent circulations of actively evolving variants of the virus. As of 2 November, a total of 8,501,503 COVID-19 cases and 218,568 deaths have been reported in African Continent, accounting for ∼3–4% of global share (1). To reduce these numbers, there is an urgent need for equitable distribution of safe and effective vaccines to the Continent. The number of SARS-CoV-2 vaccine doses deployed in countries within the African Continent is growing gradually, however, more efforts are needed to reach targets for control (2). As of the November 2021, at total of 276 million vaccines are distributed to the continent, with very smaller fraction (5.9%) of the population being fully vaccinated (3) as compared to the 50-80% in most European countries (4). Given the limited availability of vaccine and distribution capacity within the continent, characterization of the trajectory of vaccination efforts and the expected burden of SARS-CoV-2 is necessary in planning and informing the acquisition, distribution and administration of vaccines.

SARS-CoV-2 mortality and morbidity are strongly age-specific (5). This means that an important determinant of the changing burden of SARS-CoV-2 over the course of the pandemic is the changing age profile of susceptible individuals, as these represent individuals at risk of infection. The age structure of the population determines the initial number of susceptible individuals over age groups. Subsequently, age profiles of susceptibility change because of transmission resulting from age-specific contact patterns, as well as targeted vaccination programs. Given the balance of risks expected through time, most western countries have employed age-based targeting for emergency vaccine deployment, with a goal of reaching the general population once the target coverage of high-risk and older age groups is achieved (6,7). This raises two important questions. The first question is how long it will take to reach most of the high-risk population and then expanded targeting from prioritized older individuals to more general targeting. This will depend on vaccine availability and public health infrastructure to support vaccine distribution. In our analysis we further clarify a second question of how the dynamics and thus the burden of infection is likely to unfold in the intermediate term and how this will depend on country-specific demography, social contact patterns and health infrastructure.

These questions are particularly pressing on the African continent, as dose availability remains relatively low in many countries. While Africa shares 17% of the world population (8), its only approx. 3% of the world’s doses being administered as of 6 November 2021 (4). Projecting the targeting horizon for vaccination campaign in such settings, i.e. the projected horizon required to achieve a target coverage level of the high-risk groups prior to expanding to general vaccination, as well as the subsequent dynamics requires explicit consideration of the continent’s distinct demography, social mixing patterns and health infrastructure. Across many countries in Sub-Saharan Africa, a large share of the younger age classes is suggestive of a potentially higher risk of transmission among these high-contact young people. Low health system capacity (9,10) in some settings increases vulnerability to the pandemic virus. The comprehensive baseline readiness in Africa by November 2020, with an average score of 33%, is far below the minimal targeted rate of 80% (11). Such limited public health infrastructure and capacities has motivated us to contextualize the interactive effect of infections and vaccination on the evolving numbers of susceptibles-at-each-age in African settings in the near- and medium-term horizon. The in-depth analysis will inform prioritization and tailored strategies to ramp up population immunity and prevent unprecedented illness and death from COVID-19.

We illustrate with projections for diverse African LMIC demographics. To do this, we develop an age-structured Susceptible, Exposure, Infection and Recovery (SEIRS) multi-compartmental model with vaccination to allow for projections of susceptible individuals integrating age structure, mixing patterns and vaccination schedules. We employ a sequential allocation of vaccines which targets the high-risk older age groups before providing access to the general population. The age-structured SEIRS model (12) investigates how age-susceptible patterns will change with infection and vaccination according to the flow of individuals between Susceptible, Exposed, Infectious, Recovered and Re-susceptible compartments.

Assuming global mixing, we employ the simplest case for the incidence rates, which is proportional to the product of the number of susceptibles and infectives, such that virgin epidemic and thus the fraction of susceptible individuals diminish rapidly. Incorporating spatial clustering in contacts would decelerate local epidemic dynamics (13,14). To map our model to a spectrum of African-country like settings, we parametrize the model based on a broad range of demographies, social mixing and health system capacity. We frame our analyses around a best-case scenario where the health workforce is trained and available to administer vaccines, and cold chains are maintained to allow for a uniform vaccine delivery within a country. Accordingly, we use the health system capacity as the proxy of vaccine delivery capacity. The reality on ground on this best case scenario in delivering doses may differ across local settings driven by multiple factors; including spatial heterogeneities in transport network (15), vaccine hesitancy and inaccessibility to locate and thus reach the prioritized groups (16), etc.

## Methods

### Vaccine allocation

We consider a sequential allocation of vaccines to high-risk older age groups (here taken to be 65+ yrs) and the general population. Vaccination is initialized by allocating all the vaccines towards the high-risk group and subsequently made available to the entire population when 85% of the priority group is vaccinated. The logic behind this vaccination scheme is that prioritizing the high-risk at early stage to lower the burden of mortality but broadening age brackets at later stage to lower transmission and morbidity is the most common approach taken by most countries.

Assuming that *p* is the proportion of the high-risk group that will be targeted for vaccination before broad deployment and *k* is daily vaccination capacity in the population, the time length (Δ*t*) to target (referred to as the “targeting horizon”) will be 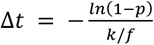, where *f* is the fraction of the high-risk group in the population. Given this, the age-stratified vaccine rate 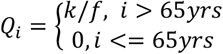 prior to expanding, and *Q*_*i*_ = *r* for all *i* after the expanded targeting. Thus, for logical consistency the rate of vaccination of the high-risk groups is reduced once the general population have access to vaccination.

We project population-wide vaccination capacity to different countries by *k*_*c*_ = *k*_0_ *HI*_*c*_, where *k*_0_ is base rate (taken as 0.001 doses per health workforce per day in the study), and *HI*_*c*_ is the index of health system capacity in country *c* (see “*Model parameterization*”). With the rates we estimate the targeting horizon for each country and present the relative targeting horizon across countries. We further integrate the estimates of age-stratified vaccination capacity in each county within a SEIRS framework to project the fraction of susceptible individuals-at-age over time. (see “*Model structure*” and “*Model simulation*”).

### Model structure

We integrate our realistic age-structured multi-compartmental model (12) with vaccination to allow for projecting dynamics of susceptible individuals under diverse scenarios of age structure and mixing patterns.

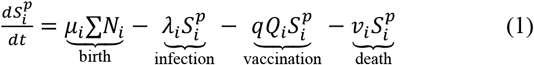

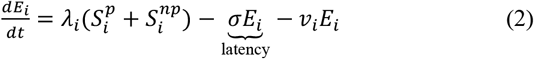

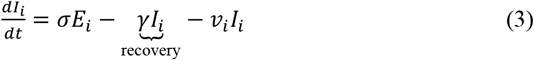

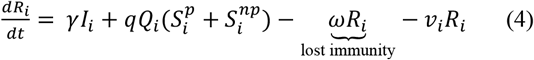

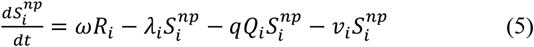

where 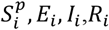 are respectively the number of primary susceptible, exposed and, infected and recovered individuals in age group *i*. The force of infection on susceptible individuals in age-class *i* at time *t* is 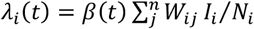, where *β*(*t*) is the rate of seasonal transmission (see “*Transition rates*”), *W*_*ij*_ is the normalized contact rate between age group *i* and *j* in the population (17,18). Age-specific vaccination capacity (*Q*_*i*_) is stratified by stage of vaccine allocation (see “*Vaccine allocation*”), with *Q*_*i*_ = *k*/*f* for the high-risk older age groups and *Q*_*i*_ = 0 for the rest of the population prior to broadening age brackets; and *Q*_*i*_ = *k* for all *i* after general dissemination; *q* is the vaccine efficacy(19). The immunity from infection and vaccination wanes with an average rate of *ω*, and the immunized individuals develop to non-primary susceptible status 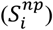.

To lay out the general framework and findings, we underpin the model with several assumptions. First, we base our assessment of age profile of susceptible individuals in the best-case scenario where we assume that health workforce is trained and cold chain is established for vaccine delivery, and that access to vaccination is equal within a country (which we acknowledge is an unrealistic assumption (15)). Additionally, we assume a homogeneous susceptibility to infection and clinical fraction over age classes. Further, we assume a temporary protection against both infections and the progression to severe disease following recovery or vaccination. Last, we assume that vaccine-induced immunity is as high as that from prior infections and thus the rate of loss of immunity for recovered and vaccinated individuals to be the same. The latter two assumptions are consistent with the emerging evidence from mass vaccination campaigns that have reported a highly effective protection against SARS-CoV-2 infections across diverse populations in real-world settings (20–22).

### Model parameterization

#### Transition rates

We calibrate the model to a medium-level transmission setting (*R*_0_ = 2.5) by using the next-generation approach (23). Here, we do not explicitly include an exact shape of the forcing function on seasonality, given that influence of specific drivers, e.g. temperature and school term-time, has not been fully identified for SARS-CoV-2. Instead, we assume a sinusoidal forcing and model the seasonal transmission rate by *β*(*t*) = *β*_0_ (1 + *β*_1_cos(2*πt*)), where *β*_0_ is the baseline rate of transmission given by *β*_0_ = *R*_0_*γ* and *β*_1_ is the strength of seasonal forcing (assumed to be 0.2 in the analysis). The average incubation period (1/*δ*) and infectious period (1/*γ*) in the analysis is taken to be 3 and 5 days, respectively (24). Given the empirical evidence that immunity does not always prevent reinfection (25,26), we assume the average immunity duration (1/*ω*) as three years for both the recovered and vaccinated individuals. The rate of natural birth (*μ*_*i*_ ) and mortality (*v*_*i*_ ) is assumed 0 for all *i* since background demographic turnover is slow relative to infection dynamics during the initial pandemic phase and the intermedium term.

#### Demographics and social mixing across ages

We selected 16 African countries across a broad range of demography, social mixing, economic status and geographic locations. These include Burkina Faso, Cameroon, Demographic Republic of Congo, Egypt, Ethiopia, Ghana, Uganda, Kenya, Liberia, Madagascar, Morocco, Mozambique, Namibia, Nigeria, South Africa and Zimbabwe (Fig 1A). For these countries, we collected age pyramids (S1 Fig) from the statistics of the United Nations (27). Additionally, we used age-dependent contacts from Prem et al. (17,18). The age profile was aggregated into 17 5-year age brackets and the contact matrices were normalized (S2 Fig) prior to model predictions.

**Fig 1.**
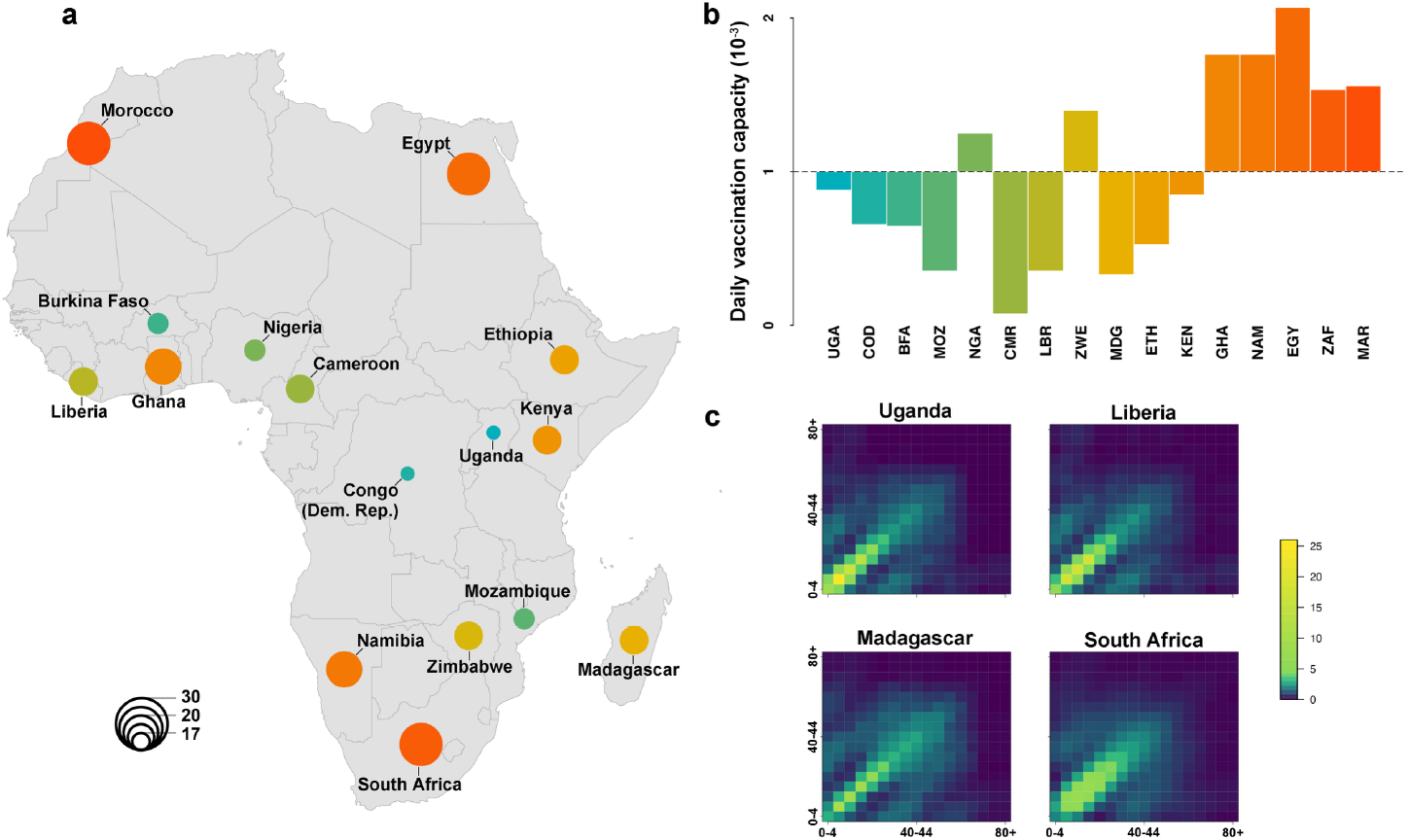
The vaccination capacity and mixing patterns in African countries. (A) 16 counties considered in the study are scaled and coloured by the median age of the population. (B) The daily vaccination capacity, i.e. the number of vaccination per health workforce per day, across counties. Countries are listed in the increasing order of the median age of the population. (C) Age-structured mixing patterns. Abbreviations: BFA: Burkina Faso, CMR: Cameroon, COD: Congo (Dem. Rep.), EGY: Egypt, ETH: Ethiopia, GHA: Ghana, KEN: Kenya, LBR: Liberia, MAR: Morocco, MDG: Madagascar, MOZ: Mozambique, NAM: Namibia, NGA: Nigeria, UGA: Uganda, ZAF: South Africa, ZWE: Zimbabwe.

#### Index of health system capacity

We collect the data on the country-wide health workforce from WHO (28) and employed this as the proxy of health system capacity. To characterize the relative capacity across countries, we define the index of health system capacity (*HI*_*c*_) as 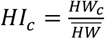, where *HW*_*c*_ is number of health workforce in country *c* and 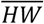 is average health workforce among all countries, such that the index fluctuates around an average of 1.

### Model simulation

We numerically integrate the model to predict disease dynamics over five years using the deSolve-package for R. Across countries with differing age structure and mixing patterns, we examine the fraction of primary and non-primary susceptible individuals over age groups at the projected horizon of expanded targeting from the high-risk to general population and at the end of year 5. For comparison, we use the age pyramids as the proxy of the age-structured fraction of susceptible individuals at the beginning of virgin epidemic where the population is fully susceptible to new pathogens. The supplement contains animations that illustrates the daily evolution of susceptibility-at-age through time.

Loss of immunity contributes to the recruitment of susceptible individuals and thus is a key in projections of disease dynamics (29). To examine its variability on our estimates, we carry out sensitivity analysis by simulating the model under scenarios with 1.5-year and 5-year duration of immunity. The fraction of susceptible individuals over age groups are estimated. The complete annotated code is available from GitHub (https://github.com/ruiyunli90/Corona-Vaccinerate).

## Results

Taking as a foundation data reflective of 16 African countries illustrates the considerable heterogeneity in age structure, vaccination capacity and mixing patterns (Fig 1, S1-S2 Fig). As a result, the targeting horizon of vaccination schedules varies with regards to two main variables: vaccination capacity based on WHO (28) and age-structure (Fig 2). Assuming a target vaccination coverage of 85% of the high-risk groups (here taken to be 65+ years of age), countries with better vaccination capacity would have a significantly shorter horizon to expand vaccination from high-risk groups to the general population. From this, Cameroon is predicted to have the longest time before general deployment due to its very low vaccination capacity as compared to other countries. The shape of age pyramids will also modulate the targeting horizon, leading to a relatively longer horizon prior to expanded targeting in countries with older population such as South Africa and Morocco.

**Fig 2.**
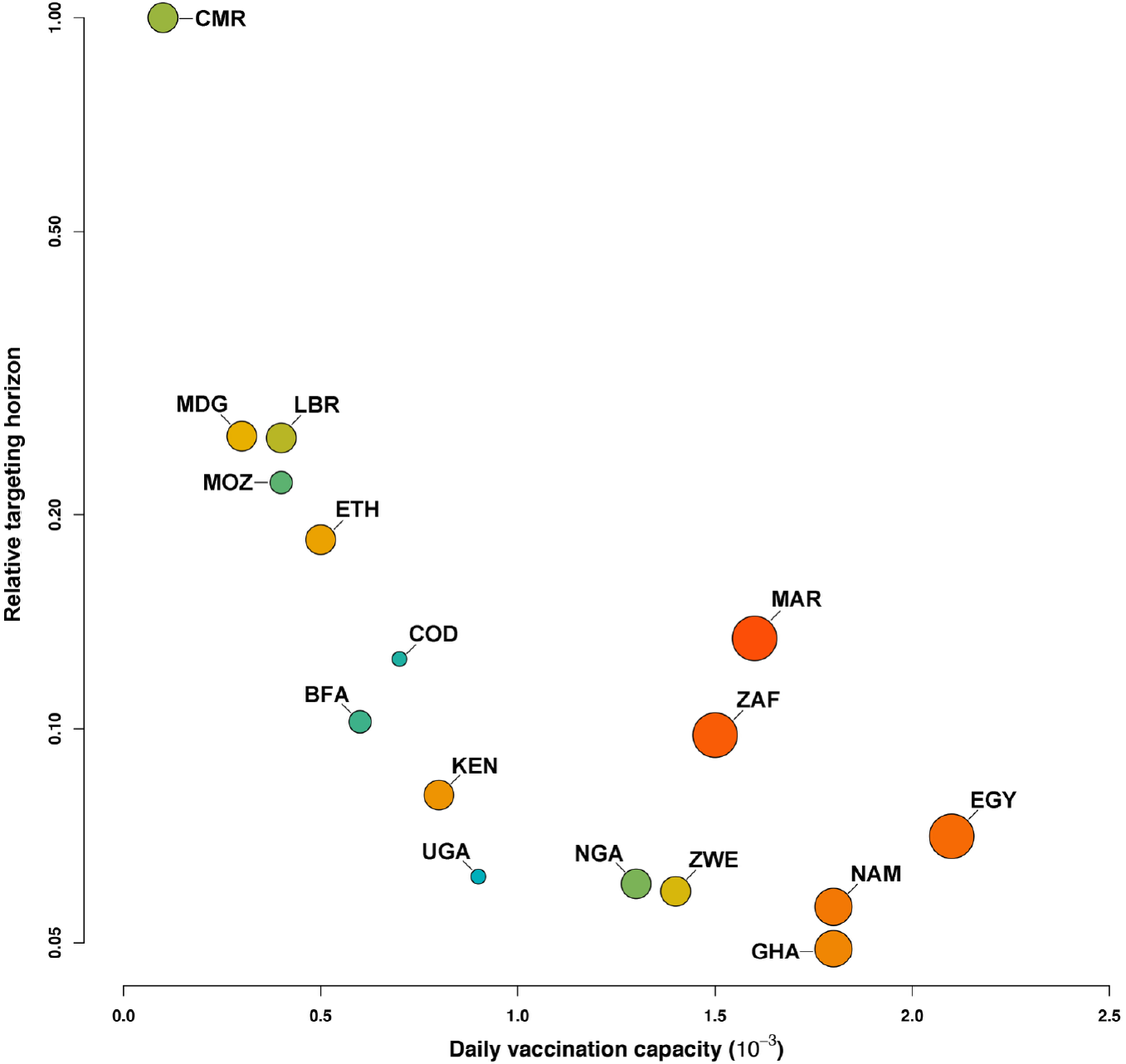
The relative targeting horizon of vaccination. Estimates of the targeting horizon, i.e. the projected horizon to make vaccination available to the general population once 85% of the old over 65 yrs is targeted, are projected to scenarios with distinct median age of the population and vaccination capacity. The targeting horizon across countries is presented on the relative scale. Circle size and colour are same with those in Fig 1. Abbreviations: BFA: Burkina Faso, CMR: Cameroon, COD: Congo (Dem. Rep.), EGY: Egypt, ETH: Ethiopia, GHA: Ghana, KEN: Kenya, LBR: Liberia, MAR: Morocco, MDG: Madagascar, MOZ: Mozambique, NAM: Namibia, NGA: Nigeria, UGA: Uganda, ZAF: South Africa, ZWE: Zimbabwe.

Once the targeting horizon is reached, our projections demonstrate that dynamics of susceptible fraction by age will change as a result of the intersection between mixing, vaccination coverage and age targeting, and loss of immunity (Fig 3, S3 Fig, S1-S16 Appendix). During the period of early emergence, infections and vaccination are dominant drivers of age profiles of susceptible individuals at the population level. By the horizon of expanding to broad deployment, vaccination will have contributed to a substantial depletion of susceptible individuals among the at-risk older age groups. By contrast, during this period the mixing patterns and thus infections-induced depletion of susceptibility among the under 65 yrs is strongly age-dependent because young people (e.g. <20 yrs in Uganda) have higher contact rates. Our model further points to the importance of loss of immunity in modulating intermediate term dynamics of age-susceptibility patterns. Under varying assumptions of immunity duration, we project a rise of non-primary susceptible individuals across age groups to the end of year 5. This projection suggests that over time the age-profile may shift as the vaccinated and recovered individuals may lose immunity and become susceptible again. These non-primary susceptible individuals, depending on country’s vaccination capacity, may subsequently be liable to be reinfected through contacts. For example, the relatively lower health system capacity of Cameroon would lead to an increase of re-infections and thus lower the fraction of non-primary susceptible individuals among the high-contact younger age classes (i.e. 5-19 yrs) in the longer term (S3 Fig). The need for booster vaccinations will depend strongly on the severity of disease following re-infection; A quantity that is currently unknown (but low for the endemic coronaviruses (26)). While the permanency of immunity to SARS-CoV-2 is still unknown, sensitivity analyses suggest that such infection-induced depletion of non-primary susceptible individuals would be universal across countries should immunity provide a shorter-term of protection (S4 Fig). A longer duration of immunity will slow the rate of reinfection (S5 Fig).

**Fig 3.**
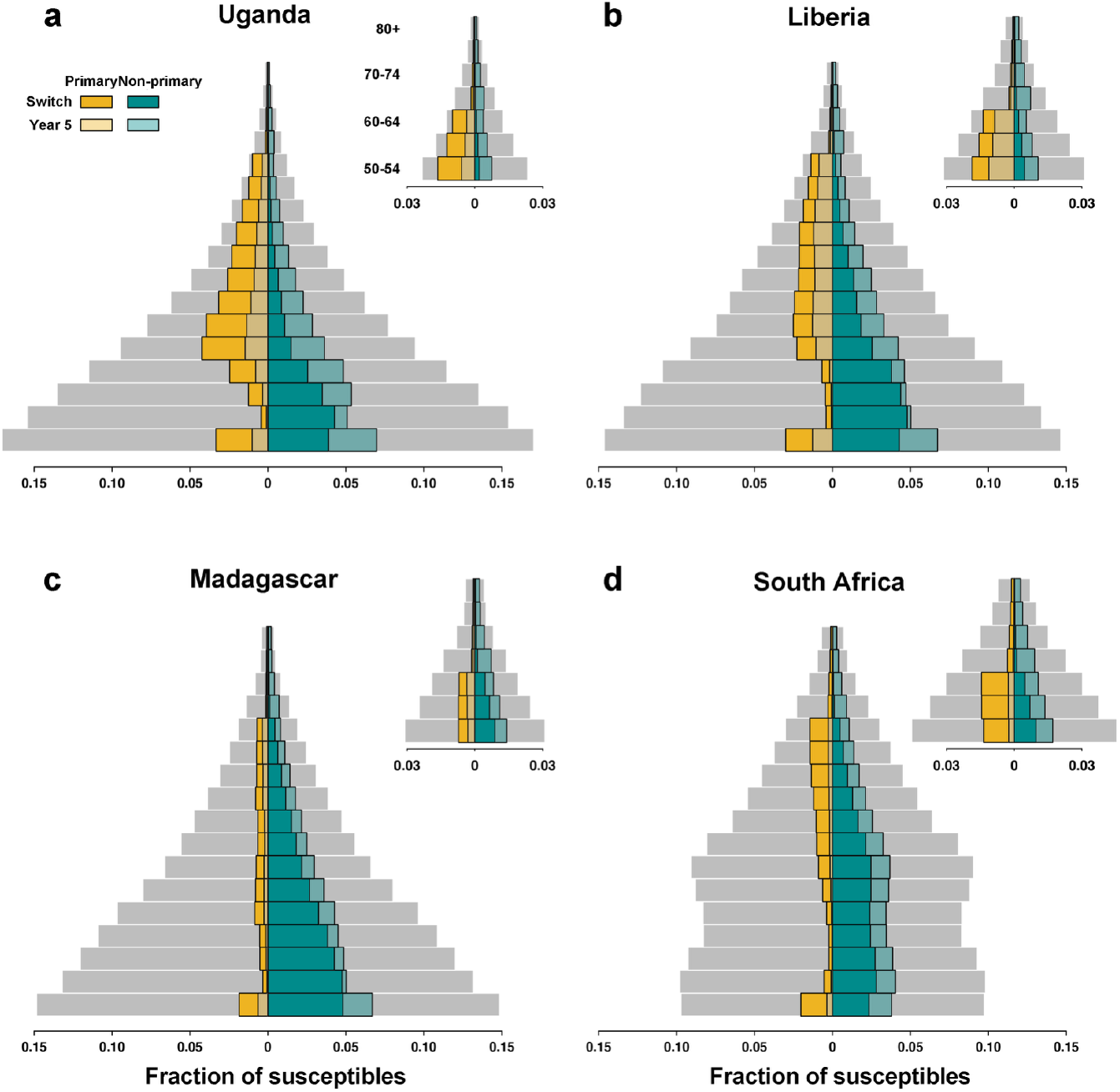
Dynamics of age-structured fraction of susceptible individuals. Age pyramids (grey bars) are used as the proxy of the age-structured fraction of susceptible individuals at the beginning of the pandemic where the population is fully susceptible to new pathogens. Assuming a three-year duration of immunity, projections of the fraction of primary (yellow bars at the left-hand side of each panel) and non-primary (green bars at the right-hand side of each panel) susceptible individuals at targeting horizon and the end of year 5 are presented. Insets are the age-profile among people over 50 yrs. Four representative countries listed in the increasing order of the median age of the population are depicted as examples to lay out the general findings.

## Discussion

Our analyses clarify the importance of considering both infection and vaccination for projections of age-profile of susceptible individuals. Future serological studies could test the predictions of the model, particularly since current vaccines should lead to a narrower antibody repertoire (mainly against the spike protein) than natural infection, allowing the two sources of antibodies to be distinguished. An important decision support tool is our simple calculation of the time-to-target cover of priority groups before general deployment depends on age-structure and vaccination capacity. In our cross-country comparison, we focused on the “relative” targeting horizon because while we consider the data on number of health care workers in each country as the determinant factor, the absolute time-to-target will be modulated by other aspects of public health infrastructure. To allow for a rapid expanding towards the target of vaccinating 60-70% of the continent’s population (approx. 800-900 million) (30), activities scaling up the supply (31–33) and improving local health systems will be critical. This demands urgent and coordinated effort for vaccine access, with regular and planned vaccine delivery aligned with the health system capacity, and to strengthen the health infrastructure including trained health workforce, planned supply delivery and improved cold chain infrastructure(34). By doing so, narrowing vaccination gap in Africa would induce knock-on benefits to global countries in the face of the emergence and spread of multiple variants of concern.

Our analysis illustrates how contact patterns and vaccination programs may be key modulators of age profile of susceptible individuals over time. Our findings demonstrate that targeted vaccination contributes to the initial depletion of susceptible individuals among the high-risk groups; while the age-specific contact patterns drives the depletion among the non-target groups. Of note, when immunity from infection and/or vaccination wanes over time, the necessity of boosting vaccination to the at-risk groups requires a deeper understanding of the severity of primary versus non-primary infections. Our projections imply that boosting vaccination would be critical to lower the burden should non-primary infections be as severe as primary ones. Our model framework is flexible in tracking both types of infections and can thus inform decision making regarding boosting vaccination once future data accumulates. Our focus on a cross-African comparison does provide important insights helping politicians and decision makers to design future vaccine-driven interventions on the African continent.

## Data Availability

All data produced in the present work are contained in the manuscript.

## Acknowledgements

We thank Dr. John N. Nkengasong (Africa CDC) for his comments on the manuscript.

## Supporting information

**S1 Fig.**
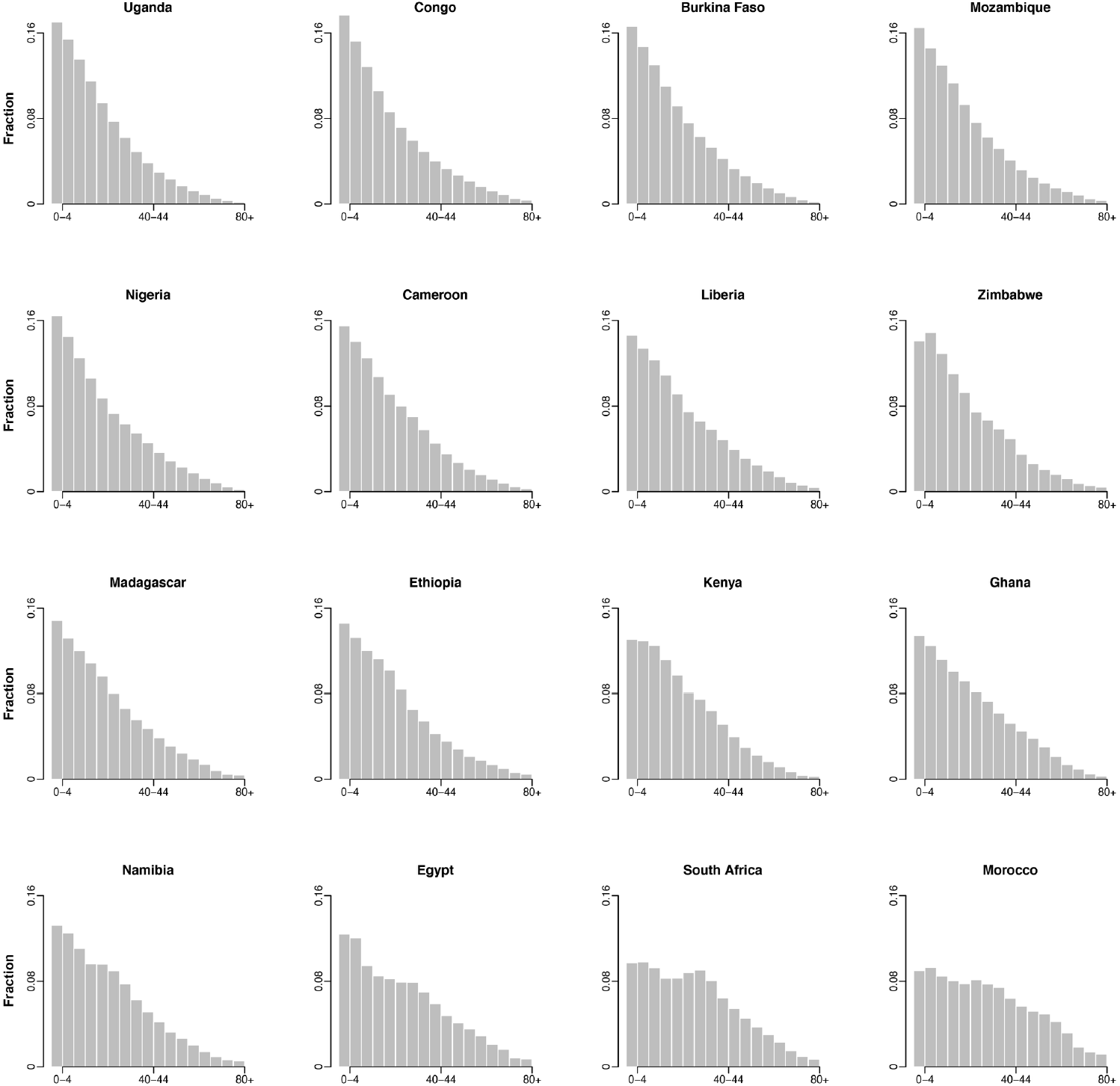
Age pyramids of 16 African countries.

**S2 Fig.**
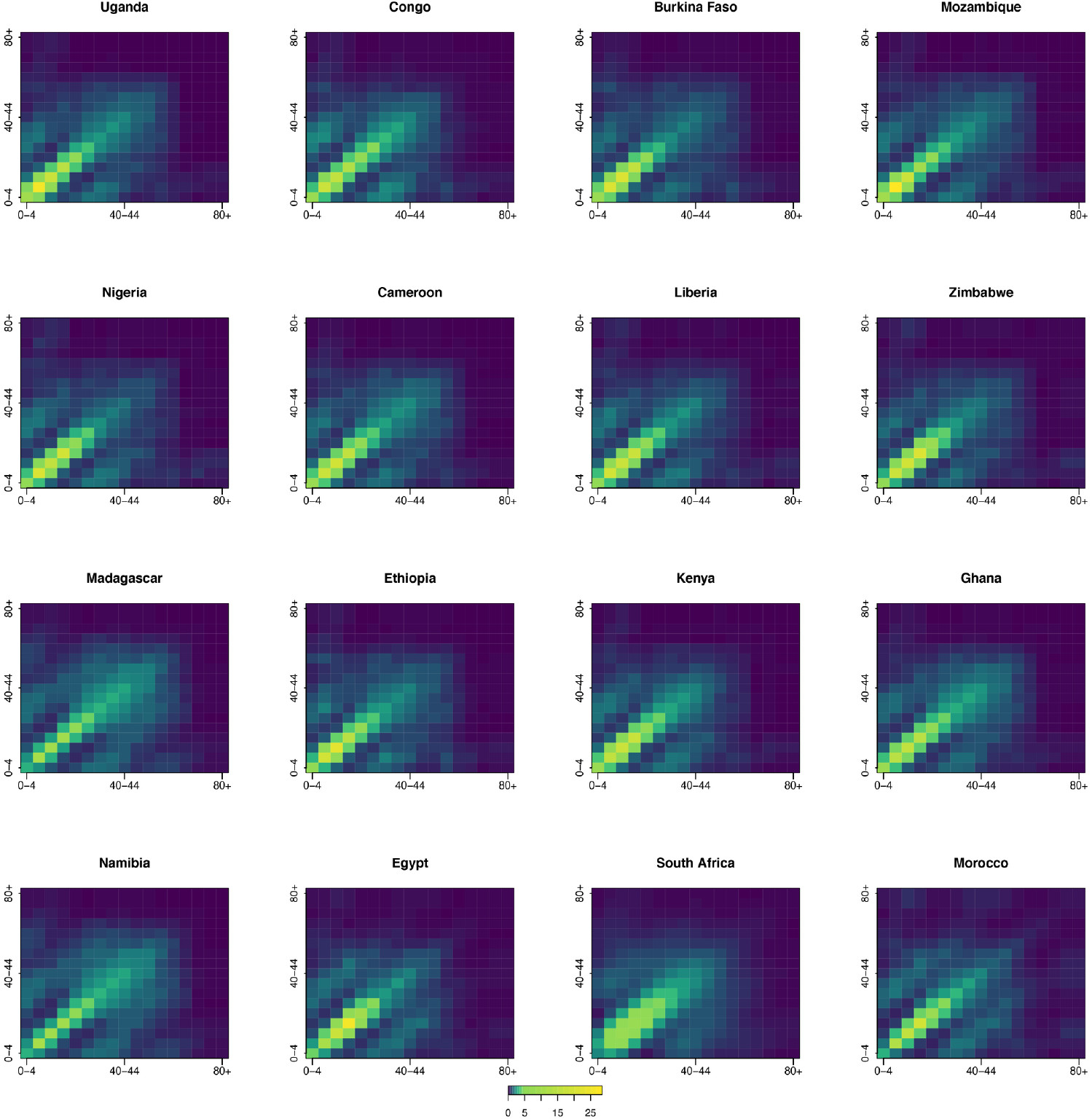
Age-structured mixing patterns of 16 African countries.

**S3 Fig.**
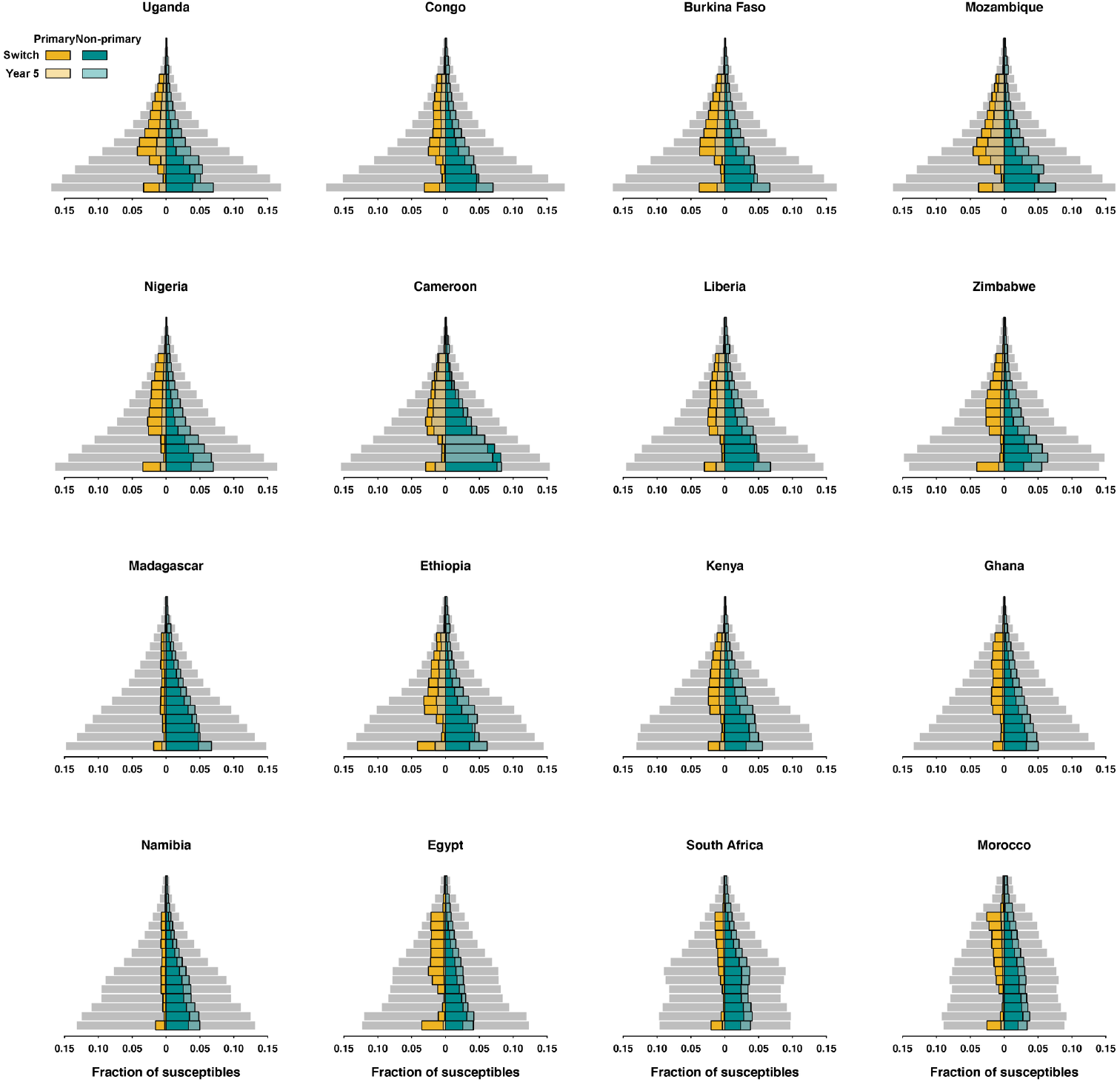
Age-structured fraction of susceptibles of 16 African countries. Same with Fig 3 but for 16 African countries in the study.

**S4 Fig.**
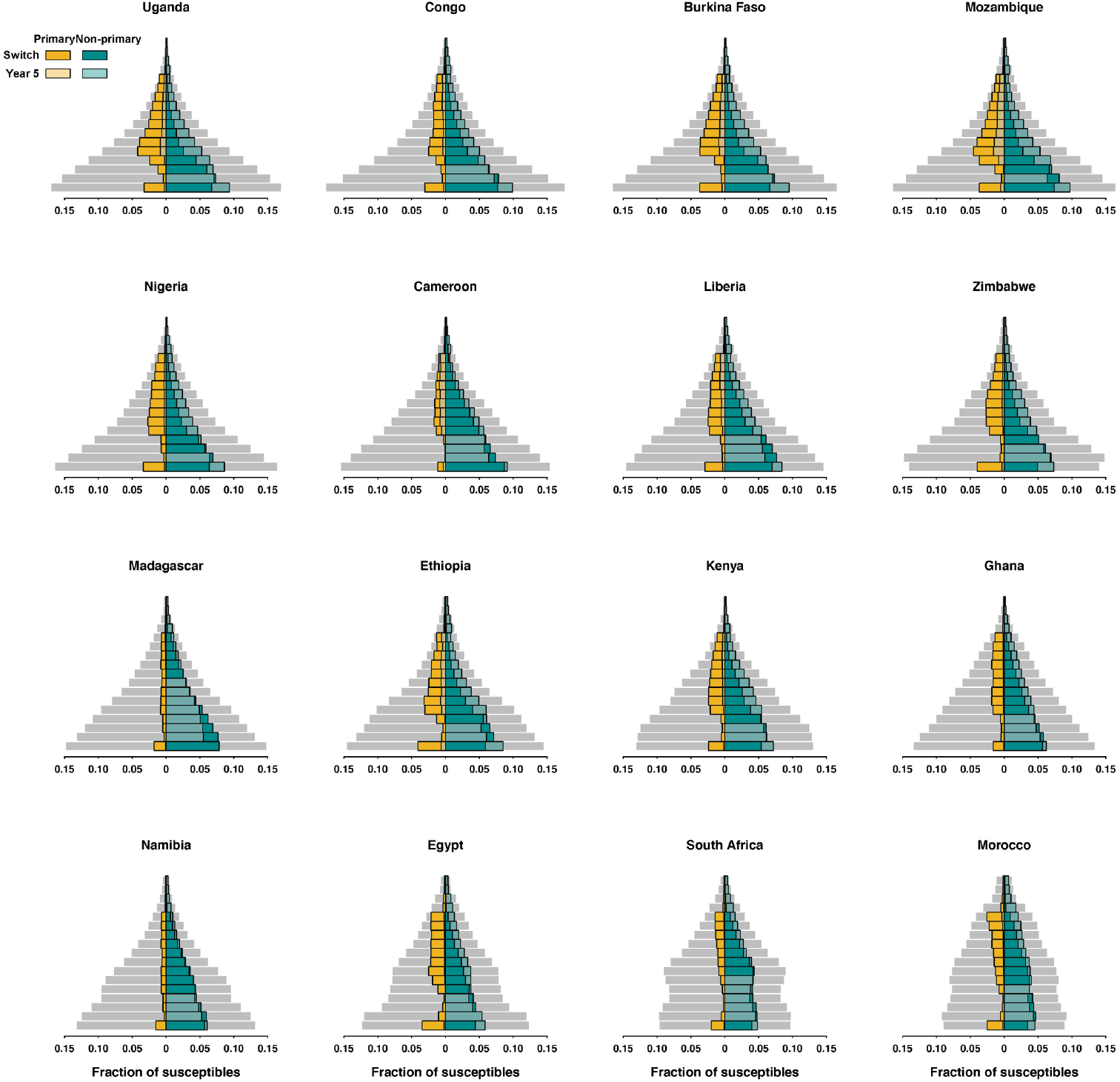
Age-structured fraction of susceptibles with alternative immune duration. Same with S3 Fig but with a 1.5-year duration of immunity.

**S5 Fig.**
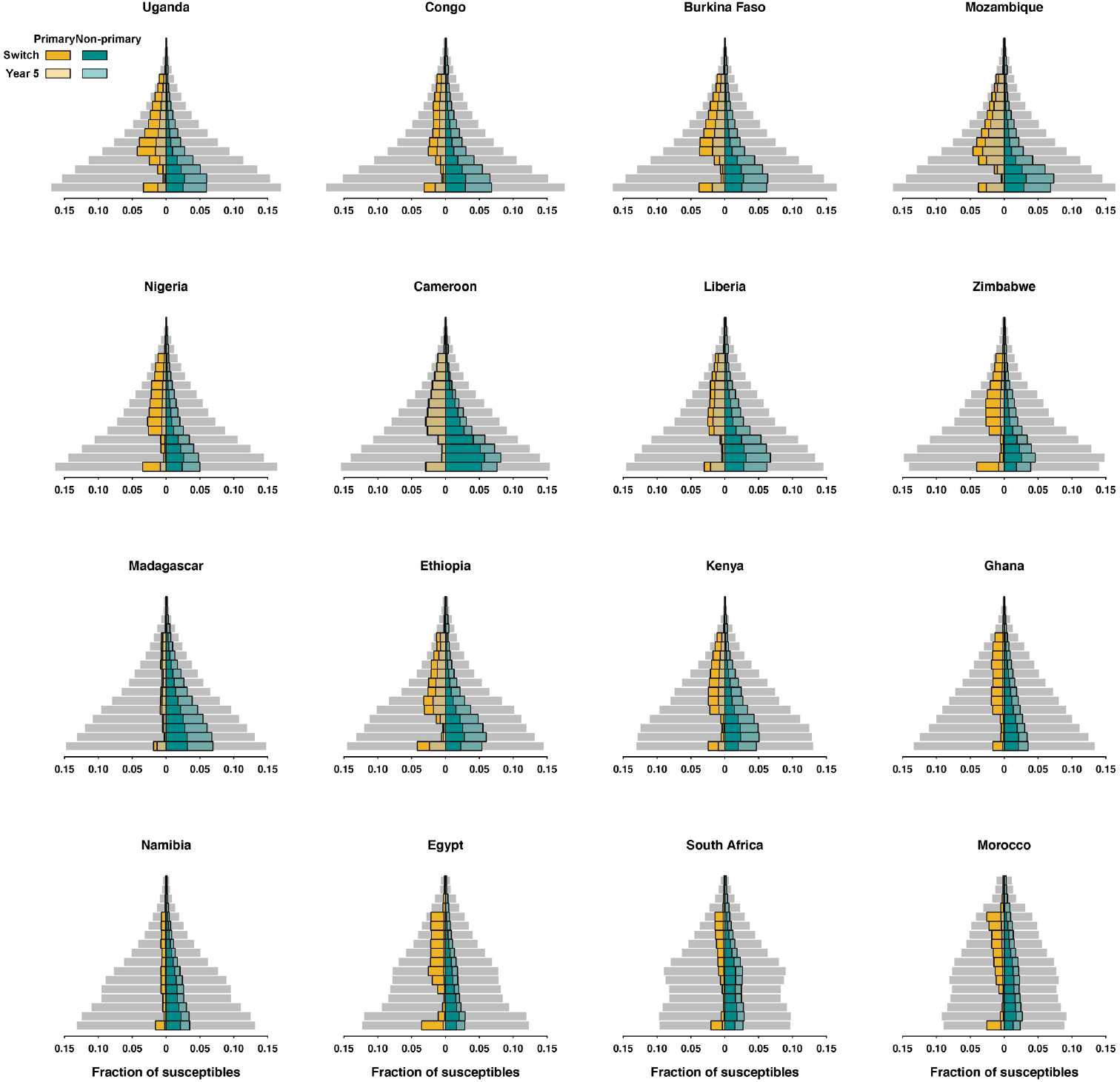
Age-structured fraction of susceptibles with alternative immune duration. Same with S3 Fig but with a 5-year duration of immunity.

**S1 Appendix. Dynamics of age-stratified susceptibles over time in Uganda**.

**S2 Appendix. Dynamics of age-stratified susceptibles over time in Congo**.

**S3 Appendix. Dynamics of age-stratified susceptibles over time in Burkina Faso**.

**S4 Appendix. Dynamics of age-stratified susceptibles over time in Mozambique**.

**S5 Appendix. Dynamics of age-stratified susceptibles over time in Nigeria**.

**S6 Appendix. Dynamics of age-stratified susceptibles over time in Cameroon**.

**S7 Appendix. Dynamics of age-stratified susceptibles over time in Liberia**.

**S8 Appendix. Dynamics of age-stratified susceptibles over time in Zinbabwe**.

**S9 Appendix. Dynamics of age-stratified susceptibles over time in Madagascar**.

**S10 Appendix. Dynamics of age-stratified susceptibles over time in Ethiopia**.

**S11 Appendix. Dynamics of age-stratified susceptibles over time in Kenya**.

**S12 Appendix. Dynamics of age-stratified susceptibles over time in Ghana**.

**S13 Appendix. Dynamics of age-stratified susceptibles over time in Uganda**.

**S14 Appendix. Dynamics of age-stratified susceptibles over time in Namibia**.

**S15 Appendix. Dynamics of age-stratified susceptibles over time in South Africa**.

**S16 Appendix. Dynamics of age-stratified susceptibles over time in Morocco**.

